# Provider and user perspectives on antenatal care delivery in KwaZulu-Natal and Limpopo, South Africa

**DOI:** 10.64898/2026.08.11.26360162

**Authors:** Danita Hingston, Thobelani Nompilo Majola, Zinhle Mtwane, Noluthando Ndlovu, Lesibana Malinga, Maanda Mudau

## Abstract

**Background:** South Africa adopted evidence-based antenatal care (ANC) frameworks to improve maternal health outcomes. However, the maternal mortality ratio remains above Sustainable Development Goal target 3.1 and routine data indicate declining ANC first-visit coverage.

**Aim:** This study sought to explore implementation gaps, barriers and facilitators in the delivery and uptake of ANC.

**Setting:** Ugu and uMzinyathi districts in KwaZulu-Natal, and Capricorn and Waterberg districts in Limpopo, South Africa.

**Methods:** A qualitative descriptive design was utilised. Semi-structured interviews were conducted with 70 purposively sampled participants, comprising 30 health system providers and 40 service users. Data were analysed thematically using NVivo®.

**Results:** Health system providers attributed declining ANC coverage to fertility decline rather than reduced access alone. Providers identified mentorship, community outreach and enhanced screening as key strengths. Implementation was constrained by staffing and equipment shortages. While service users recognised the benefits of ANC, they reported that long waiting times, negative provider attitudes and limited privacy during consultations undermined the quality of service delivery. Providers and service users linked delayed ANC initiation to financial constraints, stigma and pregnancy concealment.

**Conclusion:** Improving early ANC initiation requires an approach that addresses health system, socioeconomic and cultural barriers. Concerns about declining fertility as a driver of declining coverage warrant further investigation into coverage calculation methodologies.

**Contribution:** The study provides insights into the interconnected factors influencing ANC delivery and uptake in primary healthcare settings. It further highlights the need to consider changing fertility patterns when interpreting ANC coverage. The findings can inform targeted interventions and strengthen maternal health planning, monitoring, and service delivery.

## Introduction

Reducing maternal mortality is a public health priority in South Africa, where the maternal mortality ratio (MMR) remains above Sustainable Development Goal (SDG) target 3.1 of fewer than 70 deaths per 100,000 live births by 2030.^1^ In 2023, the country reported an MMR of 111.7 deaths per 100,000 live births.^2^ Many of the leading causes of maternal death are preventable through timely, high-quality antenatal care (ANC), making its effective implementation central to South Africa’s maternal health agenda.^1, 3^

The World Health Organization (WHO) defines ANC as care provided by skilled health professionals during pregnancy to ensure optimal maternal and newborn health outcomes.^4^ Early ANC initiation, ideally within the first trimester, enables timely detection and management of complications, promotes health education, and facilitates access to public health interventions such as HIV testing.^5, 6^ In 2016, the WHO recommended a minimum of eight ANC contacts to improve perinatal health outcomes. This recommendation emphasised not only the number of contacts, but also early initiation, continuity, and respectful standardised care.^4, 7^ South Africa adopted these recommendations in 2017 through the Basic Antenatal Care Plus (BANC+) framework within a universal, free public health system.^1^

To monitor ANC access under BANC+, South Africa tracks two key indicators defined by the National Indicator Dataset (NIDS).^8^ ANC first-visit coverage is calculated as the number of first ANC visits divided by the number of expected pregnancies. ANC first-visit before 20 weeks measures early initiation as the proportion of first ANC visits occurring before 20 weeks’ gestation. According to data from the District Health Information System (DHIS) reported in the District Health Barometer (DHB), between 2020/21 and 2024/25, ANC first-visit coverage declined from 81.7% to 61.6%, while early ANC initiation increased from 67.9% to 72.8%.^9^ Taken together, the decline in first-visit coverage and persistence of late initiation for more than one-quarter of women point to challenges in achieving universal and timely ANC coverage.

Substantial provincial variation in early ANC initiation is also evident across South Africa. In 2024/25, KwaZulu-Natal (KZN) recorded the highest proportion of women initiating ANC before 20 weeks’ gestation (78.5%), while Limpopo (LP) reported considerably lower performance (64.8%).^9^ These disparities suggest the influence of contextual factors that are not captured by routine service indicators.

Furthermore, qualitative evidence comparing ANC policy implementation and practices across provinces with contrasting performance remains limited. This study therefore aimed to explore implementation gaps, barriers and facilitators in the delivery and uptake of ANC within primary healthcare (PHC) settings in KZN and LP from the perspectives of healthcare providers and service users.

## Conceptual framework

This study was guided by the Socio-Ecological Model (SEM), which posits that health-seeking behaviours are influenced by interacting factors at the individual, interpersonal, community, organisational (health system), and policy levels.^10^ Originally developed as the Ecological Systems Theory and subsequently adapted for health promotion, the SEM provides a framework for understanding how influences across these levels shape women’s engagement with ANC services.^11, 12^ It also supports qualitative inquiry by enabling exploration of lived experiences of study participants.

In this study, the SEM is employed to examine how ANC implementation and barriers and facilitators to services operate across multiple levels, which are mapped in the findings. The individual level considers women’s knowledge, confidence, and perceptions towards ANC and other personal characteristics influencing care-seeking.^13, 14^ The interpersonal level examines the influence of women’s social relationships, including those with male partners, parents, and peers, on the uptake of ANC.^15^ The community level encompasses social norms, cultural and religious beliefs, stigma, and community outreach activities that shape access to services. ^16^ The health system level examines service delivery factors such as availability and accessibility of essential services, provider attitudes, and quality of care.^15^ Finally, the policy level considers the influence of national ANC guidelines, routine monitoring systems, and provincial and district implementation arrangements.^17^

## Research methods and design

### Study design

This study employed a qualitative descriptive design to explore gaps, barriers and facilitators in the implementation and uptake of ANC services from the perspectives of women and healthcare providers.

### Setting

The study was conducted in four purposively selected districts in two South African provinces: Ugu and uMzinyathi (KZN), and Capricorn and Waterberg (LP). Districts were selected to represent contrasting early ANC initiation contexts based on ANC booking before 20 weeks’ gestation, as reported in the DHB. In 2024/25, early booking rates were 79.9% in uMzinyathi and 77.9% in Ugu, compared with 64.6% in Capricorn and 64.3% in Waterberg.^9^

The selected districts also represent diverse geographic and socio-demographic contexts. Ugu is a district that borders the Eastern Cape province and comprises urban, peri-urban and rural communities, while uMzinyathi is predominantly rural with dispersed settlements and longer distances to health services. Capricorn district includes the provincial capital, Polokwane, and encompasses both urban and rural communities, while Waterberg is largely rural, with access to healthcare services influenced by lower population density and geographic dispersion. The study included eight facilities across rural, peri-urban and urban settings, comprising PHC clinics and community health centres. Three facilities were sampled in each KZN district and one facility in each LP district.

### Study population and sampling strategy

The study population comprised health system providers and service users. Health system providers included provincial and district maternal health managers, facility managers, and frontline healthcare workers (HCWs) involved in maternal health planning, management and service delivery. Providers were recruited using purposive sampling. Provincial and district maternal health managers, as well as facility managers, were invited via email to participate in the study. Facility managers subsequently assisted with the recruitment of HCWs by informing staff about the study and referring interested individuals to the research team.

Service users comprised pregnant women and women with infants younger than one year, aged 18–49 years. Service users were also recruited using purposive sampling at participating facilities. Facility managers introduced women attending ANC and infant immunisation services to the data collectors and provided a brief overview of the study. Women who expressed interest in participating were then given detailed study information by the data collectors and invited to participate.

### Data collection

Data collection was conducted in November and December 2025. Semi-structured key informant interviews (KIIs) were conducted with health system providers, while in-depth interviews (IDIs) were conducted with service users. Separate interview guides were developed for each participant group and were informed by the SEM. The provider guide explored health system strengths and challenges, policy implementation, routine monitoring, and perceived barriers and facilitators to ANC uptake. The service user guide focused on care-seeking decisions, experiences of ANC services, social support, and barriers and facilitators to accessing care.

Interviews with provincial and district maternal health managers were conducted by members of the research team via an online meeting platform and lasted approximately 45– 60 minutes. Trained data collectors conducted face-to-face interviews with facility managers, HCWs and service users in private rooms at participating facilities. These interviews typically lasted 30–45 minutes. All interviews were audio-recorded with participants’ written informed consent.

### Data analysis

Interviews were transcribed verbatim and verified against the recordings. NVivo® software was used to conduct thematic analysis. Data were analysed thematically using Braun and Clarke’s reflexive thematic analysis approach, which consists of five recursive phases: 1) data familiarisation, 2) coding across the dataset, 3) collation of related codes into potential themes, 4) iterative review and refinement of themes, and 5) final theme definition.^18^ Deductive codes were informed by the study aim and SEM, while inductive coding captured unanticipated aspects, with an open ‘Other’ code included for emerging factors. Themes were supported by illustrative quotations and organised to reflect both provider and service user perspectives.

Trustworthiness was strengthened through strategies addressing credibility, dependability, confirmability, and transferability.^19^ Credibility was enhanced through interviewer training, triangulation and regular multidisciplinary team discussions. Dependability was supported by a transparent and systematic analytic process guided by Braun and Clarke’s thematic analysis framework.^18, 19^ Confirmability was strengthened through an audit trail documenting methodological and analytical decisions, reflexive notes and field memos that captured observations, researcher reflections, and potential sources of bias. Transferability was supported through detailed descriptions of the study setting, participants, and health system context, enabling assessment of relevance to similar settings.

### Ethical considerations

The study was conducted in accordance with the 2024 Declaration of Helsinki.^20^ Ethical clearance was granted by the Pharma-Ethics Independent Research Ethics Committee (Ref No: 250727293). Permissions to conduct interviews were obtained from the Provincial Health Research Committees of KZN (Ref: KZ_202509_058) and LP (Ref: LP_2025-09-028), and relevant district and facility management structures. Written informed consent was obtained from all participants prior to data collection. No personal identifying information of participants was collected. Audio recordings were permanently deleted from the recording devices, and all study data were stored securely on password-protected computers accessible only to the research team.

## Results

### Health system providers

#### Socio-demographic characteristics of health systems providers

Thirty health system providers participated in the study (Table 1). Just over half of the participants were from KZN (53.3%), with the remainder from LP (46.7%). One-third of the sample were provincial and district maternal health managers (33.3%), while two-thirds were HCWs and facility managers (66.7%). Participants were predominantly female (90.0%). In terms of maternal healthcare experience, 3.3% had less than one year of experience, 20.0% had 1–5 years, 40.0% had 6–10 years, and 36.7% had more than 10 years of experience. A greater proportion of providers in KZN had more than 10 years of maternal healthcare experience compared with LP (43.8% vs 28.6%).

**Table 1:** Socio-demographic characteristics of health system providers.

|  | KZN health system providers (N=16) | LP health system providers (N=14) | Total (N=30) |
| --- | --- | --- | --- |
| Variable | n (%) | n (%) | n (%) |
| **Participant type** |  |  |  |
| Provincial Maternal Health managers | 2 (12.5) | 2 (14.3) | 4 (13.3) |
| District Maternal Health managers | 4 (25.0) | 2 (14.3) | 6 (20.0) |
| HCWs and facility managers | 10 (62.5) | 10 (71.4) | 20 (66.7) |
| **Province/district level** |  |  |  |
| Provincial | 2 (12.5) | 2 (14.3) | 4 (13.3) |
| Ugu | 7 (43.8) | - | 7 (23.3) |
| uMzinyathi | 7 (43.8) | - | 7 (23.3) |
| Capricorn | - | 6 (42.9) | 6 (20.0) |
| Waterberg | - | 6 (42.9) | 6 (20.0) |
| **Sex** |  |  |  |
| Female | 13 (81.2) | 14 (100.0) | 27 (90.0) |
| Male | 3 (18.8) | 0 (0.0) | 3 (10.0) |

| Age group |  |  |  |
| --- | --- | --- | --- |
| 25-39 | 5 (31.3) | 8 (57.1) | 13 (43.3) |
| 40-54 | 7 (43.8) | 5 (35.7) | 12 (40.0) |
| 55 years and older | 4 (25.0) | 1 (7.1) | 5 (16.7) |
| Years of experience |  |  |  |
| Less than 1 year | 0 (0.0) | 1 (7.1) | 1 (3.3) |
| 1 to 5 years | 3 (18.8) | 3 (21.4) | 6 (20.0) |
| 6 to 10 years | 6 (37.5) | 6 (42.9) | 12 (40.0) |
| More than 10 years | 7 (43.8) | 4 (28.6) | 11 (36.7) |

#### Findings from health system providers

Health system providers described factors influencing ANC implementation and access across individual, interpersonal, community, health system, and policy levels of the SEM.

#### Individual level

##### HIV-related concerns

According to providers’, fear of HIV testing discourages some women from attending ANC, while others who are aware of their HIV-positive status may be reluctant to engage with treatment services.

> *‘When they come to the clinic we have to know your HIV status. Some are afraid that they’ll test positive and then they have to start taking treatment.’* (HCW 1, Ugu, KZN)
>
> *‘They tend to delay a lot especially when they know that they have an HIV diagnosis and they’re not on treatment, they delay the first visit to the point that most just deliver at the facility.’* (District manager 1, uMzinyathi, KZN)

##### Perceptions of care leading to healthcare bypassing

Patterns of healthcare bypassing were described, particularly in Ugu district, where ANC services are sought by some women travelling from neighbouring areas of the Eastern Cape. Providers suggested that women perceive these facilities as more convenient or prefer them over facilities closer to home.

> *‘We see a lot of people coming from the Eastern Cape … the rural areas. Most of them it is people who leave the clinics near them, they prefer to come here because it is more convenient.*’ (HCW 1, Ugu, KZN)

##### Unemployment and lack of finances

Financial constraints were identified by providers as a barrier to timely ANC initiation, particularly among younger unemployed women.

> ‘*Most of them usually say that they don’t have money for transport because they are young and not working.’* (HCW 1, uMzinyathi, KZN)

#### Interpersonal level

##### Pregnancy concealment

Providers reported that adolescents and young women often conceal their pregnancies from parents or caregivers, delaying ANC initiation. Dependence on parents or caregivers for transport costs also delays care by making pregnancy disclosure necessary.

> *‘You find that teenagers hide their pregnancies from their parents because they are young so that contribute to delays.’* (District manager 1, Waterberg, LP)
>
> *‘They say, “I was scared to ask for money at home because my mother does not know that I am pregnant.”’* (HCW 1, uMzinyathi, KZN)

#### Community level

##### Community outreach activities

Community-based outreach was viewed as a key facilitator of ANC uptake. In KZN, CHWs were described as playing a central role in pregnancy identification, tracing women who missed appointments, and strengthening continuity of care between facilities and communities.

> *‘We’ve got a linkage form, it’s connected also to our community caregiver… if there’s a problem…we are missing a lady, we also communicate with them.’* (HCW 1, Ugu, KZN)

Providers from LP also highlighted use of media to encourage early ANC attendance.

> *‘Our community radio stations, we have slots where we give health education… as soon as you miss your period, you must visit your clinic.’* (Provincial Manager 2, LP)

##### Xenophobic issues

Providers highlighted xenophobia and fear of discrimination as barriers to ANC access among migrant women. Anti-immigrant sentiments within communities were perceived to discourage some women from seeking care.

> *‘Foreigners who may get discriminated against so that’s a problem for a certain proportion, especially with these organisations who are trying to stop foreigners from coming to antenatal clinics.’* (Provincial manager 2, KZN)

#### Health system level

##### Health system strengths supporting ANC

Across districts, participants identified several systemic strengths supporting ANC delivery. In KZN, midwife mentors and sub-district Maternal, Newborn, Child and Women’s Health (MNCWH) coordinators were highlighted as key sources of clinical guidance and real-time support for frontline staff.

> *‘In KZN, each district has recently employed midwife mentors… about three or four covering different clinics in the district. This intervention has potential to really improve the quality of antenatal care.’* (Provincial manager 2, KZN)
>
> *‘We are providing consistent support and an open-door policy. If they want a quick opinion on a specific patient, they are able to ask telephonically, we have MNCWH coordinators.’* (District manager 1, uMzinyathi, KZN)

In both provinces, regular professional development activities, including Essential Steps in the Management of Obstetric Emergencies (ESMOE) refresher training, were viewed as important for maintaining clinical competence.

> *‘The refresher trainings like ESMOE, we do these each year. We have a budget placed aside for training because HCWs gain knowledge.’* (Provincial manager 2, LP)

Providers also highlighted clinical initiatives aimed at strengthening ANC quality, including universal pregnancy screening for all women of reproductive age during a clinic visit. In KZN, a key clinical priority was the intensified effort to address congenital syphilis through repeated testing during ANC.

> *‘Testing for syphilis during every visit, it was not done before, but now we are doing it. It helps because there are women who test negative and when they do again, she is positive.’* (HCW 1, uMzinyathi, KZN)

##### Weaknesses affecting ANC delivery

Despite these strengths, participants highlighted gaps in the availability and maintenance of essential equipment, as illustrated by one provincial manager’s account of observing an uncalibrated blood pressure machine during a facility assessment.

> *‘They didn’t have the appropriate cuff. Scales, it’s never been calibrated. Even the equipment machine never been calibrated.’* (Provincial manager 1, KZN)

Furthermore, shortages of materials for pap smears have led to the suspension of cervical examinations in some facilities, while stockouts of consumables such as glucose strips were also reported. Limited access to on-site ultrasound services was identified as an additional challenge.

> ‘*We are struggling with doing pap smears because we don’t have the equipment.’* (HCW 2, Waterberg, LP)
>
> ‘*We have most of what we need. The only thing we currently don’t have is glucose strips. We’ve been told this is a provincial issue.’* (HCW 1, Ugu, KZN)

Delays in Emergency Medical Rescue Services (EMRS) were also reported, sometimes resulting in long waits for obstetric transfers.

> *‘At EMRS we do have a challenge when we want to transfer a person. You find that there is no available ambulance. We wait for four hours; sometimes we wait for eight hours.*’ (HCW 5, uMzinyathi, KZN)

Workforce shortages were similarly described as affecting service delivery, with participants reporting rushed consultations, disrupted first-visit services and reduced specialist support.

> *‘If we had more professional nurses or midwives it would be much more effective. We end up rushing because of queues.’* (HCW 3, Waterberg, LP)
>
> *‘We used to have advanced midwife and primary healthcare nurse in every district for the District Clinical Specialist Team. But now they have become very depleted due to financial constraints.’* (Provincial manager 2, KZN)

Weaknesses in quality assurance mechanisms were also identified, with participants reporting inconsistent implementation of routine BANC+ audits.

> *‘BANC+ has an audit system, but the challenge is that the audits are hardly taking place.’* (Provincial Provider 1, KZN)

#### Policy and monitoring level

##### Perceived trends in ANC coverage and timing

Health system providers reported that routine data suggested declining ANC first-visit coverage. However, they did not interpret this as evidence that fewer women were accessing ANC services, attributing the trend instead to declining fertility rates and limitations in how ANC coverage is calculated.

> *‘We have noticed post-COVID, decline in all maternity services… in antenatal attendance and decline in deliveries and live births*, which all contribute to a declining trend.’ (Provincial manager 1, KZN)
>
> *‘The calculations of some indicators from the DHIS are misleading. We should consider that in some provinces, fertility is declining.’* (Provincial manager 1, LP)

Participants argued that ANC first-visit coverage may not accurately reflect service utilisation, because changes in birth rates alter the estimated denominator. A KZN provider suggested that obtaining ANC attendance information among women at delivery would provide a more reliable measure of ANC uptake.

> *‘That’s a flaw in the data based on year drop in number of births. What we need to look at is at the time of birth, how many people who delivered hadn’t attended antenatal care.’* (Provincial manager 2, KZN)

##### Trends in ANC early booking

Perceptions of ANC early initiation differed between provinces. In KZN, participants described improvements in early booking.

> *‘From the look of our data, ANC early booking is going up.’* (Provincial manager 1, KZN)
>
> *‘Out of people who attend, I think over 70% attend before 20 weeks.’* (Provincial manager 2, KZN)

In contrast, LP participants identified late ANC initiation as an ongoing challenge.

> *‘Women still go for ANC, it’s just that they are not coming at the right time. Instead of them coming before 20 weeks, they come after 20 weeks and it is late.’* (Provincial manager 2, LP)

### ANC service users

#### Socio-demographic characteristics of ANC service users

Forty ANC service users participated in the study, with ten women recruited from each district. Of the participants, 70% were currently pregnant and 30% were new mothers of infants younger than one year (Table 2). Overall, 45.0% of participants were 18-24 years of age, 37.5% were aged 25–29 years, and 17.5% were aged 35 years or older. Nearly two-thirds (62.5%) had secondary education as their highest level of education, while 37.5% had tertiary education. Compared with KZN, a higher proportion of those in LP had tertiary education (55.0% vs. 20.0%) and had two or more children (50.0% vs. 25.0%).

**Table 2:** Socio-demographic characteristics of ANC service users.

|  | **KZN service users**<br>**(N=20)** | **LP service users**<br>**(N=20)** | **Total**<br>**(N=40)** |
| --- | --- | --- | --- |
| **Variable** | n (%) | n (%) | n (%) |
| **Maternal status** |  |  |  |
| Currently pregnant | 12 (60.0) | 16 (80.0) | 28 (70.0) |
| New mother | 8 (40.0) | 4 (20.0) | 12 (30.0) |
| **Age group** |  |  |  |
| 18–24 | 11 (55.0) | 7 (35.0) | 18 (45.0) |
| 25-29 | 6 (30.0) | 9 (45.0) | 15 (37.5) |
| 35 years and older | 3 (15.0) | 4 (20.0) | 7 (17.5) |
| **Race** |  |  |  |
| Black | 20 (100.0) | 19 (95.0) | 39 (97.5) |
| Coloured | 0 (0.0) | 1 (5.0) | 1 (2.5) |
| **Highest level of education** |  |  |  |
| Secondary | 16 (80.0) | 9 (45.0) | 25 (62.5) |
| Tertiary | 4 (20.0) | 11 (55.0) | 15 (37.5) |
| **Employment status** |  |  |  |
| Unemployed | 16 (80.0) | 14 (70.0) | 30 (75.0) |
| Employed | 4 (20.0) | 6 (30.0) | 10 (25.0) |
| **Relationship status** |  |  |  |
| Single | 9 (45.0) | 16 (80.0) | 25 (62.5) |
| In a relationship, unmarried | 8 (40.0) | 0 (0.0) | 8 (20.0) |
| Married | 3 (15.0) | 4 (20.0) | 7 (17.5) |
| **Number of children**<br>**(excluding current pregnancy)** |  |  |  |
| 0 | 6 (30.0) | 5 (25.0) | 11 (27.5) |
| 1 | 9 (45.0) | 5 (25.0) | 14 (35.0) |
| 2+ | 5 (25.0) | 10 (50.0) | 15 (37.5) |

#### Findings from ANC service users

Women described factors influencing ANC initiation and utilisation across individual, interpersonal, community, and health system levels.

#### Individual level

##### Knowledge and perceived benefits of ANC

Service users demonstrated awareness of the importance of ANC for monitoring the pregnancy and preventing complications. Many reported attending ANC soon after confirming the pregnancy, particularly when they perceived themselves to be at increased risk.

> *‘I bought a pregnancy test and the results were positive. The next day I went to the clinic to get checked up because I wanted to make sure my child is protected…I am a high-risk patient.’* (Participant 3, Capricorn, LP).

One participant from KZN described how screening for syphilis at her first ANC visit reinforced the importance of early attendance:

> *‘When I came for my first visit, there was syphilis, which made me realise that should I have delayed, I would have had a problem.’* (Participant 8, uMzinyathi, KZN)

##### HIV-related motivations for ANC attendance

For women living with HIV, ANC was viewed as an important pathway for accessing treatment and preventing vertical transmission.

> *‘I was encouraged by the fact that I am taking treatment for HIV.*’ (Participant 8, Ugu, KZN)
>
> *‘It is important to start attending early, here I found out that I have HIV. I know what route I should take and how I should take my medication so that the child is not easily infected.’* (Participant 10, uMzinyathi, KZN)

##### Pregnancy recognition and acceptance

Late ANC initiation was attributed to delayed recognition of pregnancy as some women reported only realising that they were pregnant months into gestation.

> *‘I did not pay attention, I only noticed when I was also vomiting, then I knew. I started when I was already far gone, this is when I was five months.’* (Participant 10, Ugu, KZN)

Some women also described struggling to come to terms with an unintended pregnancy. In these instances, reluctance to accept the pregnancy was often accompanied by efforts to conceal it from others, further delaying ANC initiation.

> *‘I was in denial when I was told I was pregnant. I forced myself [to attend] today.’* (Participant 6, Ugu, KZN)
>
> *‘Because I did not want this baby, I was hiding my pregnancy and that meant I couldn’t go to the clinic because people will see me.’* (Participant 10, Waterberg, LP)

##### Poor perceptions of clinic care

Perceptions of poor service quality were also described as deterrents to early ANC attendance. One participant attributed her delayed booking to dissatisfaction with services at her local clinic.

> *‘I started my ANC visit at six months because there is no improvement in our clinic.’* (Participant 7, Waterberg, LP)

#### Interpersonal level

##### Support from family, friends and partners

Supportive social relationships played an important role in facilitating ANC attendance. According to participants, family members, friends, and partners provided both emotional encouragement and practical support, helping them to navigate pregnancy and access care.

> *‘They [family] are very supportive, they even check my next appointment date to go to the clinic and wake me up in the morning.’* (Participant 6, Waterberg, LP)
>
> *‘I was encouraged by my friends, because I was scared to tell my mother.’* (Participant 5, uMzinyathi, KZN)

Male partners were frequently described as a source of financial support, which enabled women to attend ANC appointments and meet pregnancy-related needs.

> *‘My baby’s father is supportive, financially, he has always been there.’* (Participant 2, Capricorn, LP)

##### Economic abuse

However, not all male partners supported care-seeking. One woman described experiences of controlling and abusive partner behaviour that limited her financial autonomy and ability to access healthcare.

> *‘The father of the child was a bully, he took my money when I got paid.’* (Participant 6, Ugu, KZN)

#### Community level

##### Community stigma

Negative community attitudes, especially concerns about judgement and gossip, contributed to reluctance to seek care.

> *‘In my community, they are very judgemental… adults talk about things that aren’t even true, “certain people only know how to make babies”.’* (Participant 1, Ugu, KZN)
>
> *‘They have their own opinion due to my age.’* (Participant 9, Waterberg, LP)

##### Traditional and religious practices

Traditional and religious practices formed part of some women’s pregnancy care-seeking pathways. These practices were often used alongside ANC, although some participants described relying on them as alternatives to clinical care.

> *‘I am not used to this type of ANC, I am used to the traditional one. Receiving ANC from the traditional healer, with traditional herbs. She gives me herbs to drink, touches my stomach and administers an enema.’* (Participant 6, Ugu, KZN)
>
> *‘I went to church and they gave me spiritual water to protect me and my baby.’* (Participant 3, Waterberg, LP)

##### Role of community health workers

Notwithstanding, CHWs were described as facilitators of ANC attendance through pregnancy follow-up, and encouragement to remain engaged in care.

> *‘They’d [CHWs] ask, ‘When is the baby’s due date? Show me the card I want to see.’ They know how to encourage women.’* (Participant 10, Ugu, KZN)

#### Health system level

##### Provider conduct

Women’s interactions with HCWs varied considerably; some participants described nurses as supportive and respectful, while others reported judgemental behaviour and verbal abuse.

> *‘The nurses are very friendly and explain everything in detail to the understanding of the patient.’ (Participant 1, Waterberg, LP)*
>
> *‘They shout at you, then that one goes and tells the other one that this patient was here last year and she is here again.’ (Participant 8, Capricorn, LP)*

One participant raised concerns about privacy and confidentiality during consultations, describing interruptions while being examined.

> *‘There is a lack of privacy during check-ups as other staff members frequently enter and exit while a patient is being attended to.’* (Participant 6, Capricorn, LP)

Migrant women in Capricorn district also described communication barriers and feeling marginalised within healthcare settings.

> *‘They will talk their language sometimes and I cannot hear them.’* (Participant 3, Capricorn, Limpopo)
>
> *‘It is not comfortable, particularly for those of us who are foreigners.’* (Participant 7, Capricorn, LP)

##### Organisation of ANC services and resource availability

Participants identified several operational and resource-related challenges that affected their experiences of ANC. Long waiting times were commonly reported, with some women spending most of the day at the clinic.

> *‘What makes it difficult, is that I am going to sit in the clinic from the morning until around 2 or 3.’* (Participant 9, uMzinyathi, KZN)

Women also described disruptions in care arising from missing patient records.

> *‘Sometimes, they don’t find our files. They even tell us to come the next day.’* (Participant 8, Waterberg, LP)

Shortages of medicines and limited access to diagnostic services, particularly ultrasound examinations, were additional concerns.

> *‘Sometimes there is a shortage on medication for pregnant women due to many women attending the same facility.’* (Participant 8, Waterberg, LP)
>
> *‘They could improve by doing a scan. It would be better if it were available.’* (Participant 4, uMzinyathi, KZN)

##### MomConnect implementation

Experiences of MomConnect, a national health initiative that provides maternal health information and appointment reminders via mobile phone, indicated uneven implementation of the digital health intervention.

> *‘MomConnect you get is when you come for the first visit to open a card. It tells you when you should visit the clinic.’* (Participant 8, Ugu, KZN)
>
> *‘Last time I heard about MomConnect was when I was staying in Joburg. Here in Limpopo, I have given birth 3 times and was never told about it.’* (Participant 3, Capricorn, LP)

## Discussion

The findings indicate that ANC uptake and implementation in the selected districts are influenced by interconnected individual, socioeconomic, cultural and health system factors. Although women generally recognised the value of ANC and providers identified several implementation strengths, timely ANC initiation was hindered by financial constraints, pregnancy concealment, stigma, and concerns about service quality. At the same time, providers highlighted workforce, equipment and referral challenges that constrained implementation, while also questioning whether declining ANC first-visit coverage reflects reduced utilisation or declining fertility.

One of the most notable findings was the dual role of HIV as both a barrier and facilitator of ANC initiation. Providers frequently perceived fear of HIV testing and treatment initiation as reasons women delay attendance, whereas service users described ANC as an opportunity to access treatment and prevent vertical transmission. Similar findings have been reported elsewhere in southern Africa. In Johannesburg, HIV-related stigma discouraged ANC attendance among some women, whereas in Lesotho, knowledge of HIV status and access to prevention of mother-to-child transmission services promoted early initiation.^21, 22^ These findings suggest that HIV influences ANC utilisation through women’s perceptions and experiences: when HIV is associated with stigma, fear of diagnosis, or concerns about treatment, it may discourage care-seeking, whereas when women perceive health benefits from testing and treatment, it can motivate engagement with ANC services. However, these findings should be interpreted cautiously. Service users in this study had all engaged with ANC services and may therefore have been more likely to recognise the benefits of HIV-related care. Women who never attended ANC, possibly due to HIV-related stigma or treatment concerns, were not represented. Consequently, the extent to which HIV acts as a barrier to ANC utilisation may be underestimated.

Delayed pregnancy recognition and challenges accepting unwanted pregnancies were also important influences on ANC timing. Some women delayed care because they did not initially recognise their pregnancy, a finding supported by a Gauteng study where healthcare providers linked limited awareness of early pregnancy symptoms to late ANC presentation.^23^ This consistency reinforces the role of pregnancy awareness in shaping care-seeking behaviour. Others postponed ANC while coming to terms with an unwanted pregnancy, consistent with evidence from Sub-Saharan Africa strongly linking unwanted pregnancy to delayed ANC initiation.^24^ This suggests that late ANC initiation is not solely driven by poor knowledge but may reflect a period of uncertainty and decision-making about the pregnancy. Consequently, interventions focused only on awareness may be insufficient to address the emotional and psychological factors contributing to delayed care.

Perceptions of service quality also influenced healthcare-seeking behaviour. Providers described patterns of healthcare bypassing among patients, while some service users themselves reported delaying ANC because they perceived local services to be inadequate. Similar patterns have been reported in Ghana and South Africa, where perceived quality of care influenced bypassing of local PHC facilities.^25, 26^ These findings emphasise that availability of services does not automatically translate into utilisation; women must also perceive healthcare services as responsive and beneficial.

Financial vulnerability emerged as another barrier to early ANC uptake. Although ANC services are free at the point of care, transport expenses and unemployment continue to limit ANC initiation in Sub-Saharan Africa.^27^ These challenges were closely linked to interpersonal factors. Women frequently relied on partners, family members and friends for practical and financial support, with supportive relationships facilitating ANC attendance, as also reported in Uganda.^28^ Conversely, fear of disclosure and financial dependence delayed care-seeking. One participant also described economic abuse by her partner, highlighting how unequal power dynamics within relationships can restrict women’s autonomy and subsequent ability to access care. Similarly, a quantitative study in Mozambique found that intimate partner violence was associated with reduced ANC utilisation.^29^

Community-level influences both facilitated and hindered ANC utilisation. CHWs facilitated ANC engagement through pregnancy follow-up, referrals, and health education, according to both providers and women. This finding aligns with evidence that CHWs can improve maternal healthcare utilisation and continuity of care by serving as trusted links between communities and health facilities.^30^ Conversely, stigma and social judgement discouraged care-seeking. Women described fears of gossip, criticism and negative community perceptions, particularly surrounding adolescent pregnancies. Similar barriers have been reported in Ghana, Kenya and Malawi, where concerns about social stigma contributed to delayed ANC attendance.^31^ These findings highlight the complex role of communities in shaping ANC utilisation: while community networks can support women in accessing care, they can also create social pressures that discourage timely engagement.

Traditional and religious practices also formed part of ANC decision-making. While some women used these practices alongside formal ANC, others appeared to delay engagement with health services while seeking alternative forms of support. Consistent with the findings, a study conducted in Rwanda found that women often combine traditional and biomedical sources of care during pregnancy, suggesting that they draw on multiple forms of support that they perceive as contributing to their health and wellbeing.^32^

Factors at the health system level appeared to shape both ANC implementation and user experiences. Providers described mentorship programmes, continuous professional development and strengthened screening programmes as facilitators of implementation. This underscores the importance of policy implementation at the local level, where health systems can create an enabling environment for frontline providers to translate ANC guidelines into quality care. The emphasis on strengthened syphilis screening in KZN illustrates how such implementation can enhance service quality: providers viewed screening as a key quality improvement mechanism, while women recognised its clinical benefits. This is consistent with evidence demonstrating the benefits of repeated syphilis screening for preventing adverse maternal and neonatal outcomes.^33^ Given the high burden of asymptomatic STIs among pregnant women in South Africa, improving routine screening may represent a particularly important component of ANC quality improvement.^34^

Despite these quality improvement efforts, the findings reveal persistent implementation gaps. Workforce shortages, equipment limitations and stockouts constrained providers’ ability to consistently implement national maternal care guidelines, reflecting challenges documented in previous South African research.^35^ Delays in emergency referrals reported by providers further reflect broader weaknesses in the country’s emergency transport systems.^36^ Importantly, service user accounts illustrate how these system-level challenges are experienced in practice, contributing to long waiting times, fragmented care and limited access to diagnostic services. Long waiting times are a well-documented challenge within South Africa’s PHC system and have repeatedly been identified as a barrier to ANC utilisation.^37^

Women’s experiences of provider conduct further highlight the importance of respectful maternity care. Reports of verbal abuse, breaches of privacy and judgemental treatment indicate that quality of care extends beyond clinical services alone. These experiences can undermine trust and discourage engagement with ANC. Migrant women highlighted additional barriers related to language differences, exclusion and perceived discrimination. Similarly, providers identified xenophobia as a challenge to care access, which are barriers that have been documented in Tshwane and Johannesburg.^21, 38^

The findings also highlight inconsistency in the implementation of MomConnect. Although some women benefited from the platform, others had never been enrolled despite repeated pregnancies. MomConnect has the potential to support continuity of ANC and an early evaluation suggests that women generally value the service.^39^ However, this study indicates that its effectiveness depends largely on implementation fidelity at facility level.

At the policy and monitoring level, an important finding was providers’ interpretation of declining ANC first-visit coverage. Rather than viewing declining coverage as evidence of reduced service utilisation, many participants attributed the trend to declining fertility and limitations in the way coverage is calculated. This interpretation is plausible given evidence of declining fertility and birth rates in South Africa, driven in part by reduced adolescent fertility, delayed childbearing and increased contraceptive use.^40–43^ It is also consistent with broader literature demonstrating that maternal health coverage indicators are highly sensitive to denominator estimation methodologies.^44^

However, declining fertility may not fully explain the observed trends. It is possible that both factors are operating simultaneously, with declining fertility potentially contributing to indicator trends while some women continue to experience barriers to ANC. The qualitative design of this study does not allow these competing explanations to be tested. Nevertheless, providers’ concerns highlight the importance of critically assessing whether routine indicators accurately reflect changes in service utilisation.

The implications for policy are also significant. If declining coverage is interpreted solely as reduced ANC uptake, policymakers may incorrectly conclude that access is worsening and allocate resources based on misleading assumptions. Conversely, failing to identify genuine utilisation declines could result in missed opportunities for intervention. The suggestion by a provider that ANC attendance be assessed retrospectively at delivery may therefore provide a useful complementary measure for validating routine coverage estimates and strengthening maternal health monitoring.

Finally, differences between KZN and LP suggest that local implementation contexts may influence ANC performance. While providers in KZN reported improvements in early booking, providers in LP continued to identify delayed ANC initiation as a major concern. These observations are consistent with routine district performance data and previous research from Capricorn district.^9, 45^ Although causal explanations cannot be established, the findings suggest that implementation strategies that support early ANC initiation in higher-performing settings may offer valuable lessons for districts experiencing persistent challenges.

## Strengths and limitations of the study

This study is among the few in South Africa to apply the SEM to explore ANC delivery and utilisation across multiple levels of influence, providing a comprehensive understanding of how these factors interact to shape implementation and uptake. The inclusion of both health system providers and service users also enabled triangulation of perspectives and identification of areas of convergence and divergence. Furthermore, recruitment across districts in two provinces with differing ANC performance profiles provided insights into contextual influences and opportunities for cross-provincial and cross-district learning. Together, these strengths provide a nuanced understanding of the barriers and facilitators affecting ANC delivery in South African PHC settings.

However, several limitations should be considered when interpreting the findings. Social desirability bias may have influenced responses, particularly among service users who may have been reluctant to fully criticise services, and providers who may have downplayed system shortcomings. Furthermore, as a qualitative study, the findings provide contextual insights rather than statistically representative results and are therefore not generalisable to wider populations.

The study was conducted in two districts per province. Although these districts were purposively selected to represent a range of ANC performance, they may not reflect broader provincial contexts. Recruitment was also constrained in LP, where access permissions limited the research team to one facility per district compared with three in KZN.

All service users in the study had accessed ANC services at least once. The findings therefore reflect barriers experienced by women who eventually engaged with care and may not capture the experiences of complete non-attenders, who may face more severe or different barriers.

Finally, comparisons between districts and provinces should be interpreted cautiously. The study was not designed to establish relationships, and the findings should be viewed as context-specific experiences rather than representative of ANC implementation across either province.

## Recommendations

The study highlights the need for a systems approach to improve the timeliness, quality and equity of ANC. Priority health system interventions include reducing waiting times through increased staffing and ensuring the consistent availability of calibrated, functional equipment and essential supplies for routine maternal health services. Additionally, referral pathways should be enhanced to ensure that women requiring urgent care are not delayed by weaknesses in transport systems.

Improving respectful, person-centred care is equally critical because dignity, confidentiality and clear communication shaped whether services were perceived as supportive or discouraging. ANC improvements should prioritise respectful care alongside targeted support for adolescents, women living with HIV and migrants, who may face stigma or uncertainty when seeking care. Facility improvements should also be complemented with bolstered community-based strategies that address stigma, transport barriers, pregnancy concealment and cultural or religious concerns. Strengthening mobile health services, delivering context-specific education and engaging traditional and religious leaders can help bridge these gaps and support more equitable ANC access.

At a policy level, consideration should be given to reviewing the measurement and interpretation of the ANC first-visit coverage indicator, particularly its reliance on an estimated expected pregnancy denominator that currently does not account for declining fertility. Further assessment of the indicator’s accuracy may help strengthen monitoring and improve understanding of ANC coverage performance across districts.

Finally, further research remains important to identify women who are still excluded from care altogether to ensure the health system is responsive to local barriers, changing patterns of access, and the needs of the most vulnerable populations.

## Conclusion

This study provides an analysis of the interconnected individual, social, community, health system and policy factors shaping ANC delivery and uptake in selected PHC settings in KZN and LP. Clinical mentorship, staff training, community–facility linkages and strengthened routine screening were identified as key enablers of ANC implementation. However, persistent staffing, equipment, supply chain, and referral gaps continue to constrain service delivery. Delayed ANC initiation was associated with socio-economic barriers, stigma and health system challenges, while social support networks and access to preventive treatment facilitated early uptake. The findings underscore the need for adequate resources to support effective implementation, alongside strengthened efforts to promote respectful care and community outreach. Given providers’ concerns that declining ANC coverage may reflect changing fertility patterns, routine ANC indicators should also be reviewed to better capture service coverage and support evidence-informed planning.

## Acknowledgements

The authors are grateful to the National Department of Health for supporting this study. We also extend our sincere gratitude to the KwaZulu-Natal and Limpopo Departments of Health for their valuable support in facilitating our research within these provinces, and to all the study participants.

## Author contributions

Danita Hingston: Conceptualisation, Formal analysis, Investigation, Methodology, Project administration, Software, Writing – original draft, Visualisation, Writing – review & editing.

Thobelani Nompilo Majola: Conceptualisation, Investigation, Methodology, Data curation, Formal analysis, Software, Writing – original draft, Writing – review & editing.

Zinhle Mtwane: Conceptualisation, Investigation, Methodology, Data curation, Formal analysis, Software, Writing – original draft, Writing – review & editing.

Noluthando Ndlovu: Conceptualisation, Funding acquisition, Methodology, Supervision, Writing – review & editing.

Lesibana Malinga: Conceptualisation, Investigation, Methodology, Writing – review & editing.

Maanda Mudau: Conceptualisation, Investigation, Methodology, Supervision, Writing – review & editing.

All authors reviewed the article, contributed to the discussion of results, approved the final version for submission and publication, and take responsibility for the integrity of its findings.

## Data availability

All data supporting the findings of this study are contained within the article. Owing to the qualitative design of the study and the confidentiality agreements with participants, full interview transcripts are not publicly available. Additional information or anonymised excerpts may be obtained from the corresponding author, Danita Hingston, upon reasonable request and subject to ethical clearance and institutional approval.

